# Real-World Effectiveness of Digital Therapeutics for Hypertension Management

**DOI:** 10.1101/2024.03.03.24303639

**Authors:** Akihiro Nomura, Shuntaro Sato, Yusuke Takagi, Tomoyuki Tanigawa, Masayuki Takamura, Koichi Node, Kazuomi Kario

**Affiliations:** Department of Cardiovascular Medicine, Kanazawa University Graduate School of Medical Sciences, Kanazawa, Japan; Division of Convergence Science, Kanazawa University Graduate School of Frontier Science Initiative, Kanazawa, Japan; Frontier Institute of Tourism Sciences, Kanazawa University, Kanazawa, Japan; Noto Resilience and Revitalization Center, Kanazawa University, Kanazawa, Japan; Clinical Research Center, Nagasaki University Hospital, Nagasaki, Japan; CureApp Inc., Tokyo, Japan; Teikyo Academic Research Center, Teikyo University, Tokyo, Japan; Department of Cardiovascular Medicine, Saga University, Saga, Japan; Division of Cardiovascular Medicine, Department of Medicine, Jichi Medical University School of Medicine, Tochigi, Japan

## Abstract

**Background:** Hypertension is a major modifiable risk factor for cardiovascular disease that affects approximately 43 million people in Japan. Although lifestyle modifications are recommended as first-line interventions, their implementation within clinical settings remains challenging. Following regulatory approval of a digital therapeutic (DTx) for hypertension in Japan in 2022, we evaluated its real-world effectiveness, particularly among demographics previously excluded from clinical trials.

**Methods:** Practice-based, real-world data from patients prescribed CureApp HT between September 2022 and December 2023 were analyzed. Participants who were ≥18 years old and had at least one baseline morning blood pressure recording were included and followed for 24 weeks. The primary outcome was a change in the morning home systolic blood pressure (SBP), whereas the secondary outcomes included other blood pressure parameters and app usage metrics such as individual app engagement proportion and in-app program durations. Statistical analyses included multivariable linear regression models to evaluate between-subgroup differences while adjusting for baseline characteristics including baseline SBP levels and seasonal timing of DTx initiation.

**Results:** Among the 2038 eligible patients, mean age was 55.8 ± 11.4 years and 48.8% were female. Morning home SBP decreased from baseline at week 12 (−4.8 mmHg; 95% confidence interval [CI], −5.3 to −4.4) and week 24 (−6.0 mmHg; 95% CI, −6.6 to −5.4), with corresponding reductions in other blood pressure parameters. Consistent benefits were observed across subgroups, including older adults (≥65 years; −6.6 mmHg at week 24) and those on antihypertensive medication (−5.2 mmHg) who had previously been excluded from clinical trials. Multivariable analyses identified baseline SBP and seasonal timing of DTx initiation as determinants of treatment response, with smaller reductions having been observed with summer initiation than with spring initiation. Furthermore, good app usage metrics, including high individual app engagement and reduced in-app program durations, promoted increased SBP reductions.

**Conclusions:** Real-world evidence demonstrates that a DTx designed for individuals with hypertension effectively reduced blood pressure across diverse patient populations, with baseline SBP, seasonal factors, and patient engagement significantly influencing treatment efficacy. These findings provide important clinical insights for optimizing daily digital hypertension management.

## Introduction

Hypertension, defined as an elevation in the systolic blood pressure (SBP), has been identified as a major modifiable risk factor that greatly contributes to the global prevalence of cardiovascular illnesses.^1^ In fact, experts estimate that around 43 million people in Japan suffer from hypertension.^2^ Around 90% of all cases have essential hypertension due to the critical involvement of various lifestyle factors, such as excessive salt intake, obesity, and inactivity, in its progression.^2^ Current guidelines recommend that all hypertensive patients initially undergo lifestyle adjustments.^3^ However, the poor persistence of these lifestyle modifications and the complexity of offering individualized lifestyle guidance within the constraints of outpatient consultation times have created much difficulties.^2^ Long-term maintenance of lifestyle adjustments, particularly reduced salt intake, has been proven to be quite difficult,^4^ underscoring the need for more potent techniques of lifestyle intervention.

To address these problems, digital therapeutics (DTx) applications for hypertension have been developed with the aim of offering ongoing lifestyle advice, including diet and exercise, to enhance the effectiveness of lifestyle guidance beyond the limitations of outpatient settings.^5,6^ The recent HERB-DH1 randomized controlled trial on patients with untreated essential hypertension^7^ showed that using the DTx app (CureApp HT, CureApp, Inc., Tokyo, Japan) significantly reduced blood pressures over 24 h at home and at the office and was cost-effective.^8^ Since the approval and reimbursement of DTx for hypertension in Japan in September 2022, over a thousand patients with essential hypertension have been treated using the app. However, limited knowledge has been available regarding its effectiveness in clinical settings, especially the impact of patient age and the use of existing medications.

The current study therefore aimed to elucidate the real-world effectiveness of DTx for hypertension management by evaluating blood pressure changes among patients prescribed the DTx app for hypertension using a practice-based real-world data (RWD) cohort. Moreover, we expanded our study to encompass demographics previously excluded from the HERB-DH1 trial, specifically older adults aged 65 and above and those taking antihypertensive medication at the time of DTx prescription. This strategy allows for a thorough assessment of the DTx app’s effectiveness across a larger patient population.

## Methods

### Overview

Deidentified practice-based RWD for CureApp HT were used to evaluate blood pressure trends in consecutive patients over a period of up to 6 months (**Figure 1**). Our group safely stores these data in accordance with health information privacy guidelines. Our study protocol was approved by the Medical

**Figure 1.**
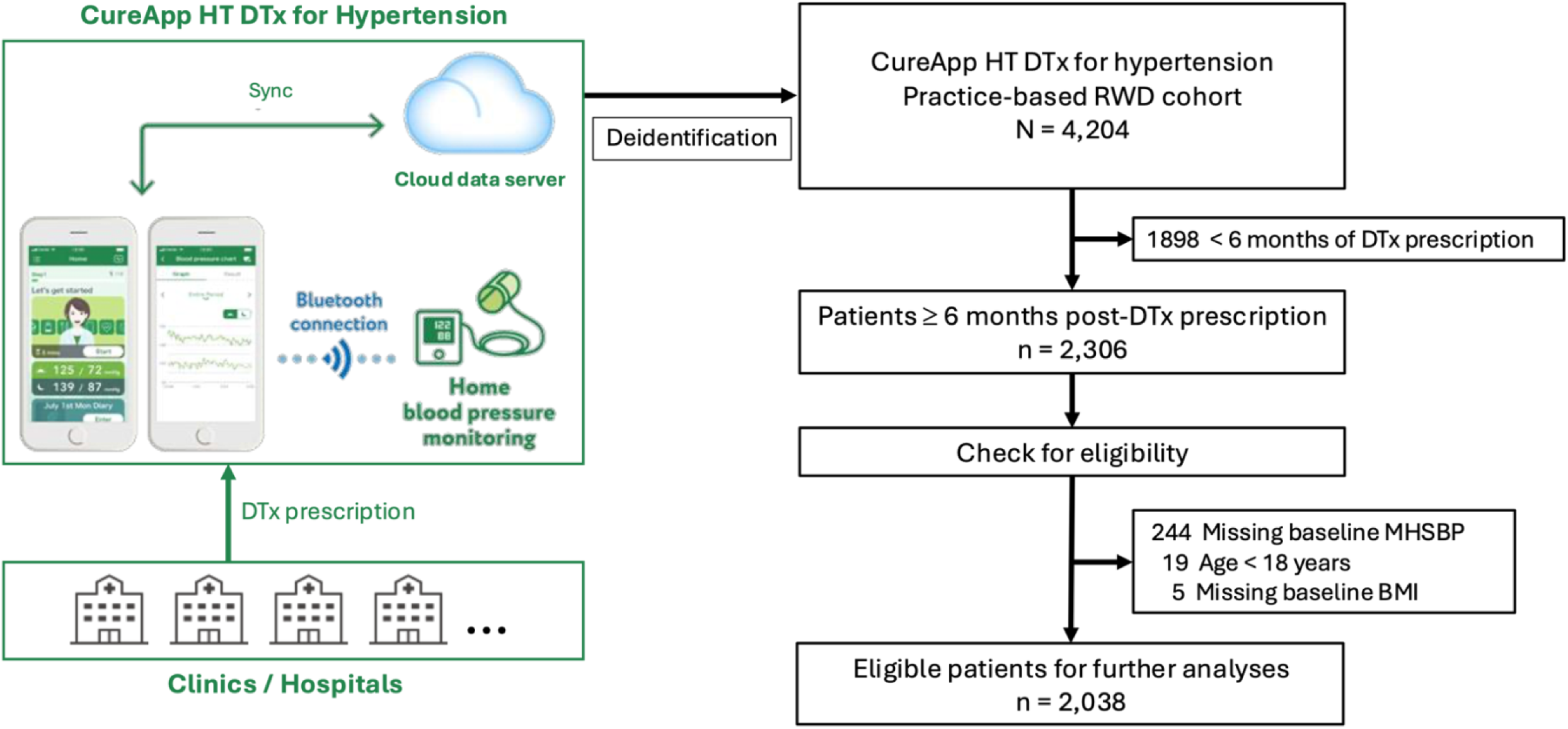
Schematic of this study. Abbreviation: DTx, digital therapeutics.

Ethics Committee of Kanazawa University and was registered with the UMIN Clinical Trials Registry (UMIN ID: UMIN000052169).

### CureApp HT

In 2022, CureApp HT, a DTx app for hypertension management, received regulatory approval and insurance coverage in Japan. This novel therapeutic modality integrates mobile app technology into outpatient hypertension care. Detailed information about the DTx app has been presented previously.^5,7,9^ In brief, this app provides continuous support for hypertension management through lifestyle modifications between clinic visits^5^ and aims to optimize nonpharmacological interventions as recommended by existing guidelines^3,10,11^ by educating patients on hypertension and promoting six evidence-based lifestyle modifications, namely salt reduction, weight management, exercise, alcohol moderation, sleep improvement, and stress management.^5^ The 6-month program consists of three phases: education about hypertension management (Step 1), guided lifestyle interventions (Step 2), and self-directed goal setting for behavioral maintenance (Step 3).^5,7^

### Criteria for inclusion and exclusion

Patients aged ≥18 years included in the CureApp HT practice-based RWD cohort were enrolled. Eligible participants were prescribed CureApp HT between September 2022 and December 2023 and required to have at least one recorded morning blood pressure during week 1 (defined as the baseline) for subsequent analyses. Patients who lacked baseline body mass index (BMI) data or showed no app usage following the prescription were excluded. All participants were followed for 24 weeks after the prescription, corresponding to the redemption period for DTx in Japan (**Figure 1**).

### Outcomes

The primary outcome was changes in home SBP from baseline to weeks 12 and 24. Secondary outcomes included changes in morning diastolic blood pressure (DBP), evening SBP, and DBP from baseline to weeks 12 and 24. The overall DTx retention rate was defined as the study-wide percentage of patients who entered their blood pressure values into the app at least once during the specified week (week 12 or 24). We also evaluated the proportions at which each hypertension guideline-recommended blood pressure threshold was achieved [(1) SBP <135 mmHg and DBP <85 mmHg based on the 2023 guidelines by the European Society of Hypertension (ESH)^11^; (2) SBP <130 mmHg and DBP <80 mmHg based on the 2017 guidelines by the American College of Cardiology (ACC) and American Heart Association (AHA)^12^; and (3) SBP <125 mmHg and DBP <75 mmHg based on the 2019 guidelines by the Japanese Society of Hypertension (JSH) ^3^] at weeks 12 and 24.

Additionally, the following app usage metrics were obtained: individual app engagement proportion, duration in the Step 1 program, and duration in the Step 2 program. Individual app engagement proportion was calculated as the percentage of days on which blood pressure was measured and input by the patient from day 1 to their last day of app usage. Step 1 phase duration was defined as the number of days from the start of app usage to the beginning of Step 2, whereas Step 2 phase duration was defined as the interval between the start of Step 2 and the start of Step 3.

### Practice-based real-world data extraction

As of September 2024, deidentified baseline profiles and blood pressure readings for all candidate patients were obtained from the cohort database. The prescription date was considered as day 1. Data were censored on the final day of blood pressure measurement or at the conclusion of follow-up (168 days, equivalent to 24 weeks). Weekly average blood pressure measurements were obtained using a 7-day arithmetic mean of each blood pressure type from weeks 2 to 24. The baseline SBP for week 1 was calculated by averaging the 6-day SBP readings from days 2 to 7, omitting measurements on day 1 as previously mentioned.^13^ Abnormal values, identified as BMI <10 or >100 and weekly average SBP of <60 or >240 mmHg, were omitted and designated as not applicable. The database did not impute missing or not applicable values. In the RWD cohort, medication usage was defined based on the app user’s declaration of “the use of medication” up until July 26, 2023, and “the use of antihypertensive medication” subsequent to July 27, 2023, as stated on the questionnaire within the DTx app.

### Statistical analysis

For baseline characteristics, continuous variables were presented as means with standard deviations or medians with interquartile ranges, depending on the data distribution, whereas categorical variables were reported as the numbers with proportions. The primary and secondary endpoints were examined using the mean with 95% confidence intervals (CI) based on the *t-distribution*.

Subgroup analyses for changes in home morning SBP from baseline to weeks 12 and 24 were conducted based on age (<65 or ≥65 years), sex (male or female), BMI (<25 or ≥25 kg/m^2^), salt check-sheet score (<14 or ≥14 points), medication status (yes, no, or unknown), baseline morning home SBP (<120, 120–130, 130–140, 140–150, or ≥150 mmHg), and season of DTx initiation (spring, March to May; summer, June to August; autumn, September to November; or winter, December to February).^14^ During subgroup analyses, between-subgroup differences were evaluated using multivariable linear regression models, with each model adjusting for all baseline characteristics (age, sex, BMI, salt check-sheet score, medication status, baseline morning home SBP category, and season of DTx initiation) except the variable under investigation. Additionally, Pearson’s correlation coefficients were calculated to evaluate relationships between morning home SBP at baseline and those at weeks 12 and 24.

For analyses of app usage metrics, we categorized individual app engagement proportion into a binary outcome (good or poor), with the threshold for “good” being set at 71.4%. This aligns with the reimbursement criterion for the use of DTx in hypertension management in Japan, which requires blood pressure readings to be taken at least five times per week (equivalent to 5 out of 7 days, or 71.4%).

Between-group differences were assessed based on the following thresholds: (1) individual app engagement proportion (≥71.4% vs. <71.4%); (2) duration in Step 1 (≥30 vs. <30 days); and (3) duration in Step 2 (≥60 and <60 days). Between-group differences were also evaluated using multivariable linear regression models to adjust for age, sex, BMI, salt check-sheet score, medication status, baseline morning home SBP, and season of DTx initiation. Data visualization and analyses were performed using R version 4.4.2, which was developed by the R Foundation for Statistical Computing in Vienna, Austria, and included the use of R packages including “dplyr,” “tidyverse,” “car,” “ggplot2,” “grid,” and “forestploter.”

## Results

The baseline characteristics of eligible patients are displayed in **Table 1**. The median follow-up duration was 168 days (interquartile range [IQR], 137–168 days). The sample comprised 48.8% females, with a mean age of 55.8 ± 11.4 years (range, 18–92 years). Baseline home blood pressure readings were 133.5 ± 13.5 mmHg (morning SBP), 86.2 ± 10.3 mmHg (morning DBP), 129.4 ± 14.0 mmHg (evening SBP), and 81.6 ± 11.1 (evening DBP). Regarding the seasonal distribution of DTx prescription, the number of patients enrolled during autumn and winter was twice that enrolled during spring and summer, reflecting the study enrollment period (from September 2022 to December 2023).

**Table 1.** Baseline characteristics of patients in the practice-based real-world data cohort.

|  | **Total** |
| --- | --- |
| **N** | 2,038 |
| **Age**, mean ± SD | 55.8 ± 11.4 |
| **Female**, n (%) | 995 (48.8) |
| **Salt check-sheet score**, median [IQR] | 12 [9–16] |
| **BMI**, median [IQR] | 24.5 [22.2–27.6] |
| **Antihypertensive medication**, n (%) |  |
| Yes | 823 (40.4) |
| No | 514 (25.2) |
| Unknown | 701 (34.4) |
| **Baseline blood pressure (mmHg)**, mean ± SD | |
| Morning home SBP | 133.5 ± 13.5 |
| Morning home DBP | 86.2 ± 10.3 |
| Evening home SBP | 129.4 ± 14.0 |
| Evening home DBP | 81.6 ± 11.1 |
| **Baseline morning home SBP category**, n (%) |  |
| <120 mmHg | 288 (14.1) |
| 120–130 mmHg | 561 (27.5) |
| 130–140 mmHg | 626 (30.7) |
| 140–150 mmHg | 327 (16.0) |
| >150 mmHg | 236 (11.6) |
| **Seasonal distribution of DTx prescription**, n (%) |  |
| Spring (March to May) | 361 (17.7) |
| Summer (June to August) | 345 (16.9) |
| Autumn (September to November) | 626 (30.7) |
| Winter (December to February) | 706 (34.6) |

Abbreviations: BMI, body mass index; DBP, diastolic blood pressure; IQR, interquartile range; SD, standard deviation; RWD, real-world data; SBP, systolic blood pressure.

### Changes in blood pressure from baseline

For our primary analysis, we first evaluated changes in morning home SBP from baseline in the practice-based RWD group. Changes in morning home SBP from baseline were −4.8 mmHg (CI, −5.3 to −4.4) and −6.0 mmHg (CI, −6.6 to −5.4) at weeks 12 and 24 (**Figure 2**). We also assessed variations in morning home DBP, evening home SBP, and DBP (**Figures S1–S3**). At week 24, changes in each morning home DBP, evening home SBP, and DBP were −4.3 mmHg (CI, −4.7 to −3.8), −5.7 mmHg (CI, −6.4 to −5.1), and −4.1 mmHg (CI, −4.6 to −3.6), respectively. Furthermore, the overall DTx retention rates were 80.7% (CI, 78.8–82.6) and 65.9% (CI, 63.4–68.4) at weeks 12 and 24.

**Figure 2.**
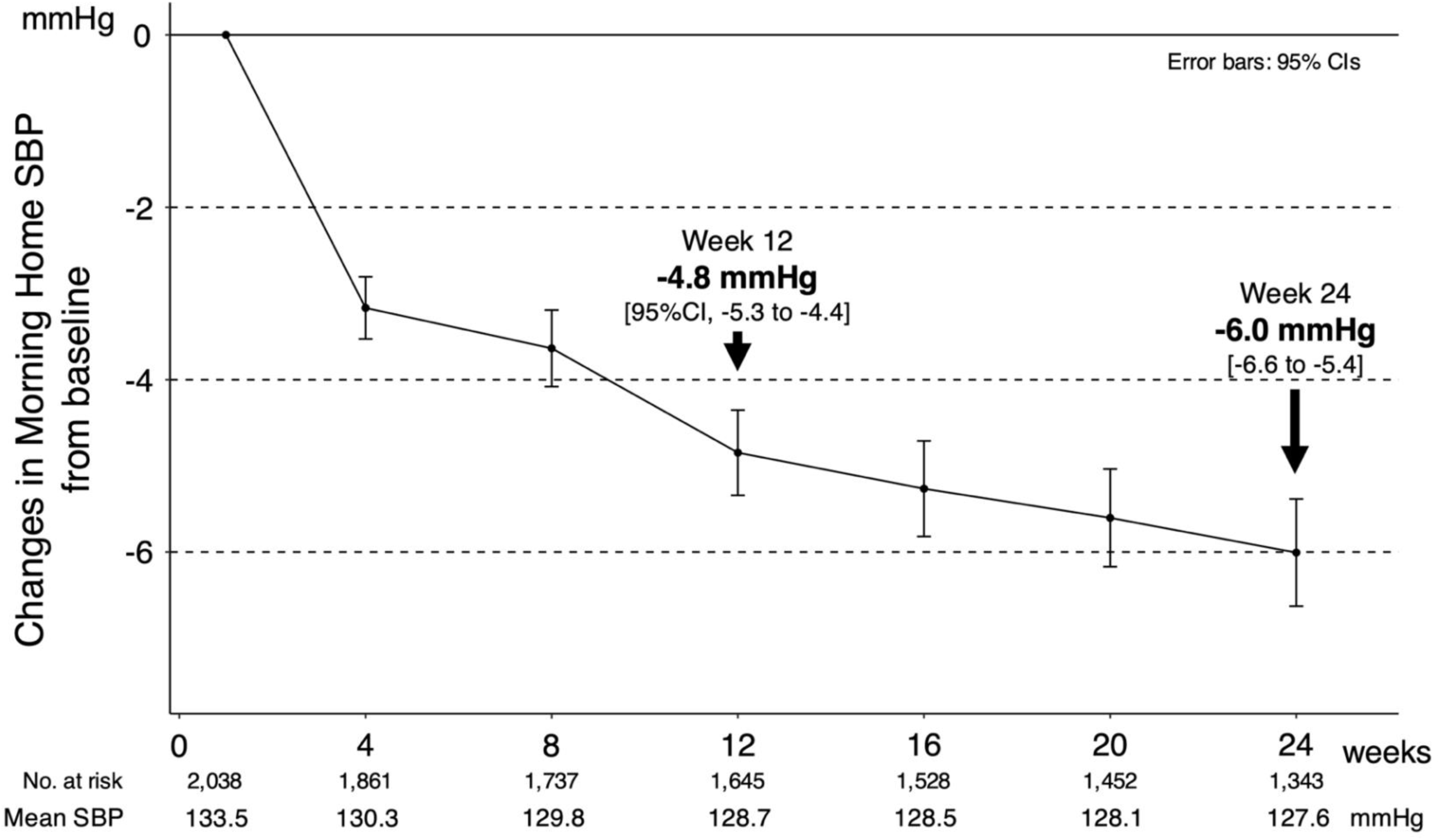
Changes in morning home systolic blood pressure from baseline among the entire study cohort. Abbreviations: CI, confidence interval; SBP, systolic blood pressure.

Additionally, we assessed achievement proportions based on each guideline-recommended blood pressure threshold (**Figure S4**). At week 24, the achievement proportions based on ESH 2023, ACC/AHA 2017, and JSH 2019 were 56.4%, 33.8%, and 15.9% for morning home blood pressure and 73.5%, 56.8%, and 33.1% for evening home blood pressure.

### Variations in morning home SBP reduction across different subgroups

Next, we assessed morning home SBP variations across the subgroups at week 12 (**Figure 3A)** and 24 **(Figures 3B and S5–S9**). Consistent SBP reduction was observed across other subgroups, particularly among patients aged ≥65 years and those on antihypertensive medication upon DTx initiation at both weeks 12 and 24. These findings demonstrate therapeutic benefits in two populations previously excluded from the HERB-DH1 trial: (1) patients aged ≥65 years (change from baseline at week 24, −6.6 mmHg [CI, −7.9 to −5.2]; adjusted difference vs. <65 years, 0.18 [CI, −1.1 to 1.5]) and (2) those on medication at baseline (−5.2 mmHg [CI, −6.2 to −4.2; adjusted difference vs. no medication, 0.74 [CI, −0.57 to 2.0]). Notably, baseline morning home SBP and seasonal timing of DTx prescription were associated with SBP changes from baseline at both weeks 12 (**Figure 3A**) and 24 (**Figure 3B**). The magnitude of the SBP reduction was strongly influenced by baseline SBP levels (**Figure 4A**), with higher baseline values associated with greater reductions in DTx users (**Figure S10**). Moderate correlations were observed between baseline morning home SBP measurements and those measured at week 12 (correlation coefficient, r = 0.676) and week 24 (r = 0.544) (**Figure S11**). Regarding seasonal variation, patients initiating the DTx in summer showed smaller SBP reduction at 24 weeks than did those initiating the DTx in spring (reference), indicating temperature-related seasonal effects (**Figures 4B and S12**).

**Figure 3.**
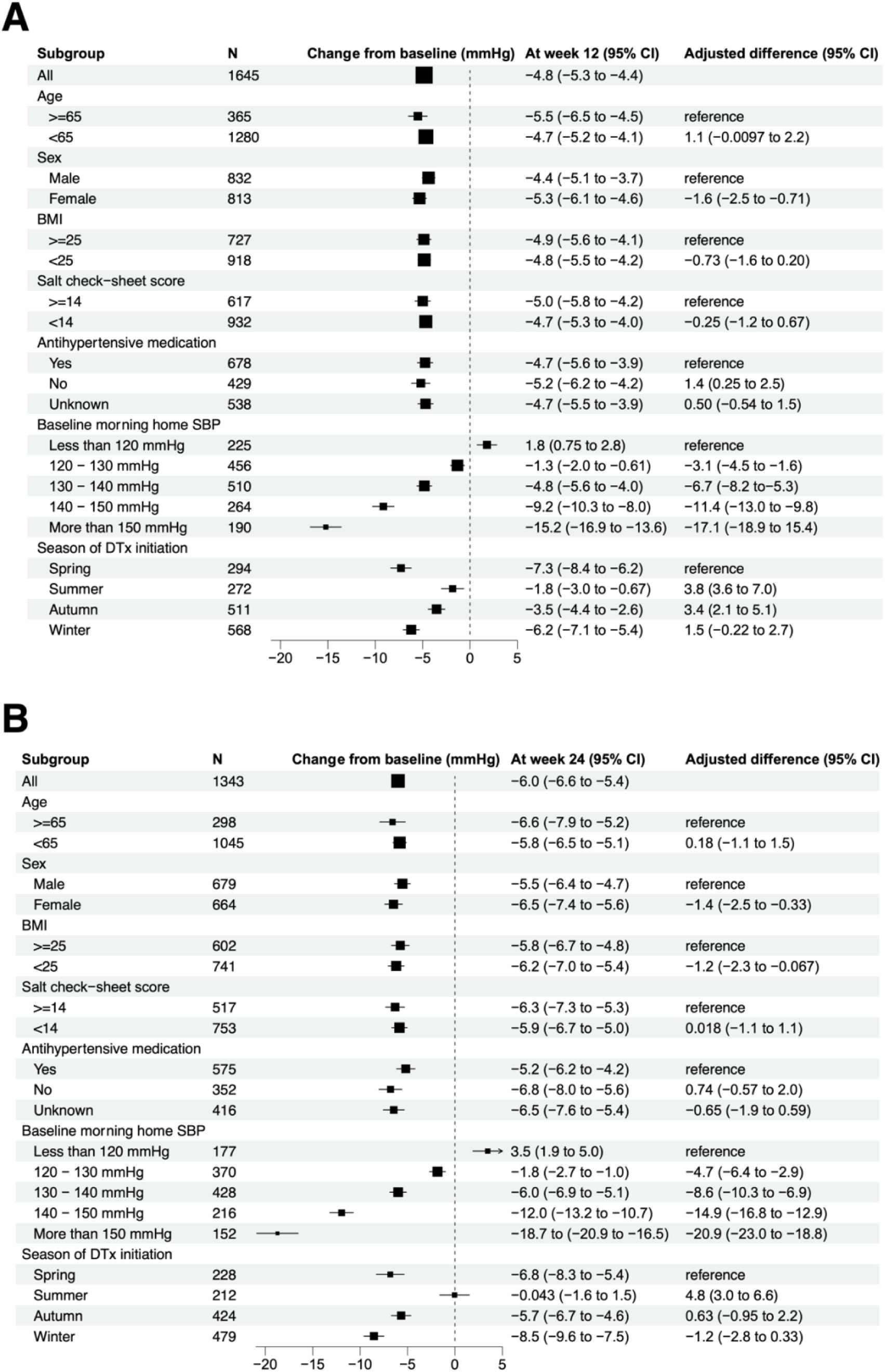
Subgroup changes in morning home systolic blood pressure (SBP) at weeks 12 (A) and 24 (B). Morning home SBP reductions were consistent across subgroups except for baseline morning home SBP and seasonal distribution of digital therapeutics (DTx) prescription. Our analysis revealed the practical benefits of DTx for managing hypertension in patients aged 65 years or older and those receiving medication at baseline, which were populations specifically excluded in the previous HERB-DH1 trial.

**Figure 4.**
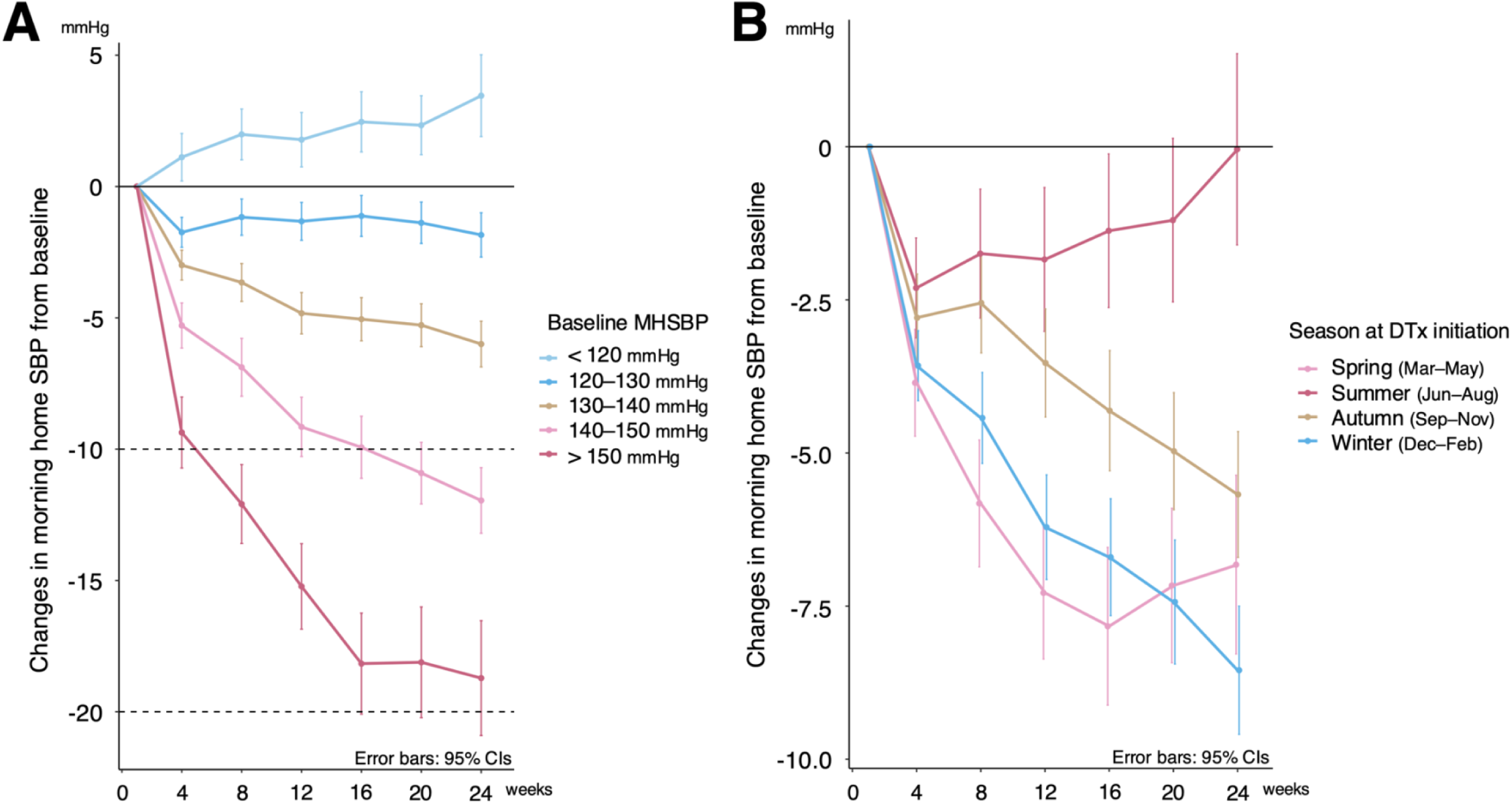
Changes in morning home systolic blood pressure (SBP) from baseline according to baseline morning home SBP (A) and seasonal variation of digital therapeutics (DTx) prescription (B). Abbreviation: MHSBP, morning home systolic blood pressure.

### Association between app usage metrics and blood pressure reduction

Finally, we examined the relationship between the three app usage metrics (individual app engagement proportion, Step 1 duration, and Step 2 duration) and SBP changes at weeks 12 and 24 (**Figure 5**). Notably, patients with high individual app engagement (≥71.4%) achieved greater SBP reductions than did those with low engagement (<71.4%) at both week 12 (adjusted difference, −1.7 mmHg; CI, −3.0 to −0.43) and week 24 (−3.4 mmHg; CI, −5.1 to −1.6). Regarding program progression, a shorter Step 1 duration (<30 days) was associated with greater SBP reduction than was a longer Step 1 duration (≥30 days) at week 12 (1.9 mmHg; CI, 0.85 to 3.0), although no such difference was observed at week 24 (0.36 mmHg; CI, -0.95 to 1.7). Similarly, patients who completed Step 2 in less than 60 days demonstrated greater SBP reduction than did those requiring ≥60 days, with such a difference being significant at week 24 (1.8 mmHg; CI, 0.66 to 3.0).

**Figure 5.**
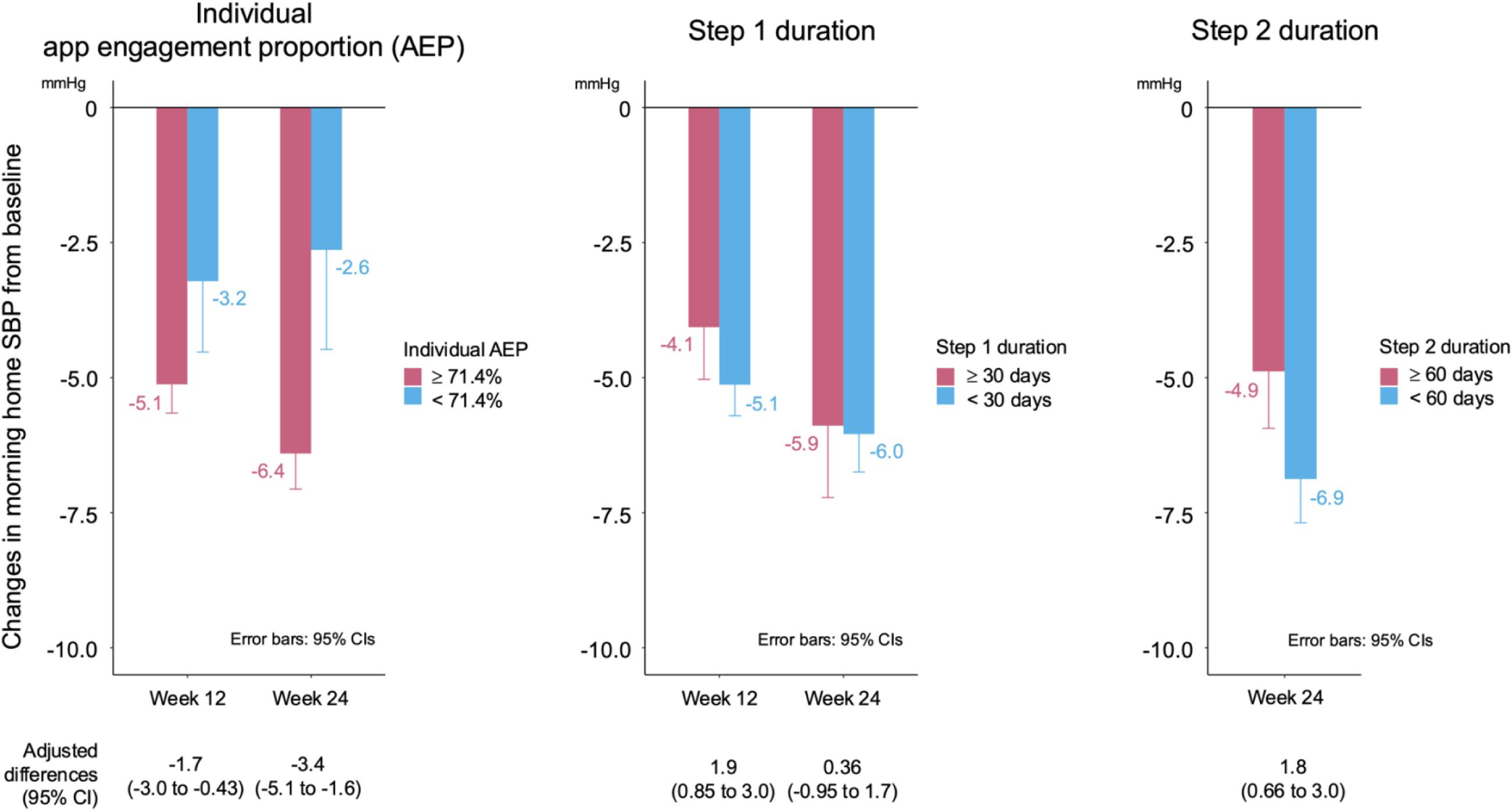
Changes in morning home systolic blood pressure from baseline according to app usage-related parameters: individual app engagement proportion (A), Step 1 duration (B), and Step 2 duration (C).

## Discussion

The current study used a practice-based RWD cohort to evaluate blood pressure changes in patients prescribed the DTx app for hypertension. Our results demonstrated reductions in morning home SBP from baseline at both week 12 (−4.8 mmHg) and week 24 (−6.0 mmHg). Moreover, the DTx app conferred realistic benefits across diverse hypertensive patient subgroups, with its effects being notable among older adults (≥65 years) and those on medication—demographics who had been previously excluded from the HERB-DH1 randomized controlled trial. Importantly, baseline morning home SBP levels and the season at which DTx was initiated significantly influenced the magnitude of blood pressure reduction at both assessment timepoints. Furthermore, higher individual app engagement proportion and more efficient progression through the Step 1 and Step 2 programs were associated with greater reductions in morning home SBP.

This research offers several important findings. First, our findings show that DTx certainly lowers blood pressure in real-world scenarios, supporting the results of a recent meta-analysis by Boima et al., which showed that digital interventions promoted a significant reduction in SBP.^15^ Moreover, our subgroup analysis demonstrated that the DTx app induced consistent morning home SBP reductions across subgroups, especially among patients over 65 years old and those on medication, emphasizing its effectiveness in populations previously excluded from the HERB-DH1 trial.^7^ This analysis demonstrates the value of RWD in revealing the effectiveness of the DTx app in populations not included at the clinical trial stage, addressing clinical questions difficult to address through randomized controlled trials alone (e.g., the correlation between glucose status and baseline blood pressure on cardiovascular diseases from the Japanese nationwide claims-based database^16^ or first-line treatment choices and medication adherence for hypertension management from TriNetX electronic medical records in the United States^17^).

Notably, baseline SBP and the seasonal timing of DTx initiation were identified as important determinants of treatment response, with baseline SBP already at optimal levels and summer initiation promoting smaller blood pressure reductions. Stratification of baseline morning home SBP values revealed a baseline SBP-dependent linear drop in SBP, consistent with the findings of Gazit et al. on the efficacy of a smartphone-based hypertension self-management program.^18^ Despite correlations between baseline morning home SBP and subsequent measurements, the confluence of equality and regression lines around the optimal SBP threshold of 120 mmHg^2,3,12^ demonstrates the potential of comprehensive lifestyle improvements encouraged by the DTx app to promote optimal SBP ranges. For example, the PREMIER study clearly demonstrated that total lifestyle adjustment impacted blood pressure in patients with grade 1 hypertension whose average SBP approached 120 mmHg after a 6-month intervention.^19^ Conversely, patients with a baseline home morning SBP below 120 mmHg showed an increase in SBP at 12 and 24 weeks, which trended toward the 120 mmHg mark. This paradoxical effect had been reported in a previous study showing that SBP gradually approached 120 mmHg over a 3-year period in patients using smartphone-based hypertension self-management, beginning with normal blood pressure (SBP <120 mmHg) at baseline.^18^ Given that one of the primary mechanisms through which DTx achieves blood pressure reduction is dietary sodium reduction,^7^ our findings align with those presented in previous research by He et al., which revealed that the blood pressure-lowering effects of salt reduction were substantially lower in normotensive individuals than in hypertensive patients.^20^ This physiological phenomenon may explain the diminishing effectiveness of the DTx intervention as the patients’ blood pressure approaches the normotensive range. Hence, further investigations into the mechanism by which the DTx app influences patients already in the ideal SBP range, as well as studies evaluating the long-term effectiveness of the DTx app on cardiovascular outcomes beyond blood pressure control, are imperative.

Regarding seasonal variation, the timing of DTx initiation was associated with morning home SBP changes at both weeks 12 and 24, with SBP being 3.8–4.8 mmHg higher with summer initiation than with spring initiation. Blood pressure exhibits well-established seasonal fluctuations in response to environmental temperature as a consequence of physiological thermoregulation mechanisms.^21^ Home morning SBP typically registers 5–6 mmHg lower in summer compared to winter,^21–23^ which aligns with our findings. Given an overall DTx-mediated reduction in morning home SBP of −4.8 and −6.0 mmHg at weeks 12 and 24, respectively, one can reasonably assume that the intervention effect for patients with hypertension who initiated the DTx during summer was essentially neutral due to the counterbalancing effect of this inherent seasonal variation in blood pressure.

Over 80% of the DTx users continued with the app through week 12, with two-thirds maintaining usage for the full 24-week period. Individual app engagement proportion could be one of the important variables for both lifestyle modification and app efficacy.^18,24,25^ Although the overall DTx retention rates were lower than those observed in well-conducted randomized controlled trials,^7^ they were consistent with those reported in previous digital intervention studies,^26^ suggesting that the results obtained in the current study were quite favorable. Furthermore, higher individual app engagement proportion and more efficient progression through the Step 1 and Step 2 programs were associated with greater reductions in morning home SBP. These results consistently highlighted the importance of app engagement and expeditious completion of specific learning modules for lifestyle modification, which likely reflects higher motivation for hypertension control, in achieving desirable blood pressure-lowering effects among DTx users.

Blood pressure control proportions among DTx app users according to the hypertension guidelines were encouraging, although some room for improvement is present. In particular, our study found control proportions of 56.4%, 33.8%, and 15.9% for morning home blood pressure and 73.5%, 56.8%, and 33.1% for evening home blood pressure based on ESH 2023, ACC/AHA 2017, and JSH2019, respectively. In this RWD cohort, hypertensive patients demonstrated reasonably good control compared to participants of the earlier HERB-DH1 trial, who were primarily poorly controlled, with all patients having a baseline morning home SBP exceeding 135 mmHg. The blood pressure-lowering effects of the DTx app seem to plateau at around a home SBP level of 120 mmHg, which is clearly lower than the target thresholds suggested by the three hypertension management guidelines (<135/85 mmHg by ESH 2023,^11^ <130/80 by ACC/AHA 2017,^12^ <125/75 by JSH 2019^3^). Moreover, the J-HOP study, a recent prospective study on home blood pressure management, showed the potential long-term benefits of stringent home SBP control below 125/75 mmHg.^27^ Nevertheless, the control levels in morning home blood pressure achieved in the current study were suboptimal, with threshold achievement proportions of 46%, 29%, and 15% for ESH 2023, ACC/AHA 2017, and JSH 2019, respectively.^28^ Although hypertension management through the DTx app showed promise for long-term cardiovascular benefits in our practice-based RWD cohort, these results highlight the urgent need for additional research to achieve the strict control objectives (<125/75 mmHg) established by the JSH2019 guidelines.

The strength of our study lies in its innovative evaluation of the practical effects of an approved and reimbursed DTx app for hypertension within a considerably large cohort. Nevertheless, certain limitations exist, including the absence of a control group who did not use the DTx app. Moreover, individual app engagement proportions might have been overestimated due to differences between the latest blood pressure reading and the actual last app use. Additionally, compared to the previous pivotal study (i.e., the HERB-DH1 trial), our study included a larger proportion of patients with normal or elevated blood pressure due to their continued use of antihypertensive medications, which may have attenuated the effect of the DTx app. Nevertheless, subgroup analyses based on medication status demonstrated that the DTx app was effective for hypertension management was regardless of whether patients were taking antihypertensive medications. Before July 27, 2023, medication usage responses may have been inaccurately recorded given that the medication confirmation item in the app questionnaire did not explicitly specify antihypertensive medications. Some patients might reduce or discontinue medication in favor of using the DTx app, although this could not be certainly established from the database alone.

In conclusion, real-world evidence obtained in the current study demonstrates that a DTx app for hypertension management effectively reduces blood pressure across diverse patient populations in clinical practice, particularly benefiting older adults and those on medication, expanding its therapeutic potential beyond previously studied demographics. The significant influence of baseline SBP, seasonal factors, and patient engagement on treatment outcomes provides crucial insights for optimizing DTx implementation in various clinical contexts. Future research should focus on enhancing patient engagement and program adherence as key modifiable factors for improved efficacy, ultimately supporting the broader integration of hypertension management into daily life with DTx.

### Contributions

AN, YT, and TT contributed to the conception and design of the study. YT and TT conducted data extraction and deidentification. AN and SS contributed to the data cleaning and statistical analyses. All authors were involved in writing the manuscript, interpreting the study findings, critically evaluating the manuscript, and approving the final form for submission.

### Declaration of interests

AN and SS received honoraria from CureApp, Inc. AN also received research grants from CureApp, Inc. YT is a CureApp employee and may own stock. TT is one of the CureApp’s board members and may own stock. KN and KK received advisory board fees and honoraria from CureApp, Inc. MT has nothing to disclose.

### Data sharing

Direct requests for access to the data underpinning this study can be made to the corresponding author. The data provider will jointly assess such requests.

## Data Availability

This study was supported by CureApp, Inc. (Tokyo, Japan).

## Acknowledgments

We would like to express our gratitude to all patients and staff who participated in this study, especially to Mitsuharu Aga, Yumi Hirayama, Fumi Hisaki, Hiroki Irisuna, and Kohta Satake for productive discussions regarding this study. We also thank Drs. Yusuke Ohya and Katsuyuki Miura for providing productive comments on the study results. This study was supported by CureApp, Inc. (Tokyo, Japan). In addition, we would like to thank Enago (www.enago.jp) for the manuscript review and editing support.

During the preparation of this manuscript, Claude 3.5 and 3.7 Sonnet (Anthropic PBC, San Francisco, USA) were used to supporting English language editing. After using this tool, we reviewed and edited generated contents as needed and take full responsibility for the contents of the publication.

## Notes

### Author Declarations

Institutional Review Board of Kanazawa University's Medical Ethics Committee

### Summary of Updates

This revised manuscript presents an increased total number of enrolled subjects for the study and incorporates additional analytical results, thereby enabling a more thorough examination.

