## Supplemental material for "Real-World Effectiveness of Digital Therapeutics for Hypertension Management"

Akihiro Nomura, Shuntaro Sato, Yusuke Takagi, Tomoyuki Tanigawa,
Masayuki Takamura, Koichi Node, Kazuomi Kario

**Supplementary Figure S1. Changes in morning home DBP from baseline**

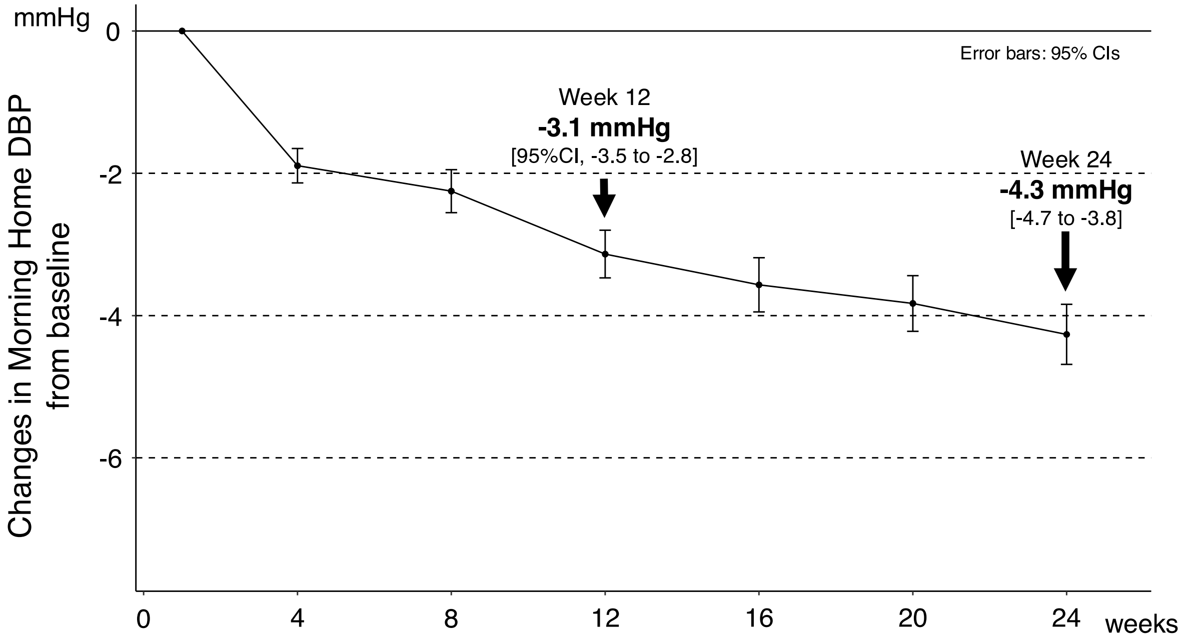


| | | | | | | | | |
|---|---|---|---|---|---|---|---|---|
| No. at risk | 2,038 | 1,861 | 1,737 | 1,645 | 1,528 | 1,452 | 1,343 | |
| **Mean DBP** | **86.2** | **84.5** | **84.1** | **83.2** | **83.1** | **82.6** | **82.3** | **mmHg** |

Abbreviations: CI, confidence interval; DBP, diastolic blood pressure.

**Supplementary Figure S2. Changes in evening home SBP from baseline**

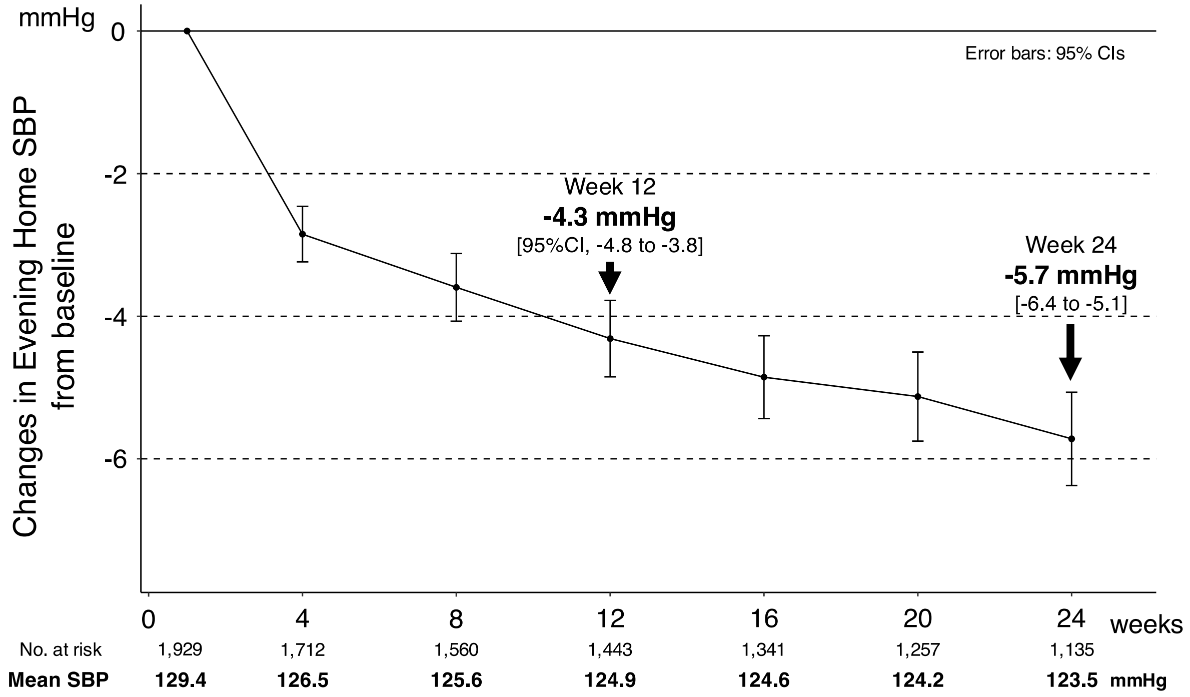


Abbreviations: SBP, systolic blood pressure.

**Supplementary Figure S3. Changes in evening home DBP from baseline**

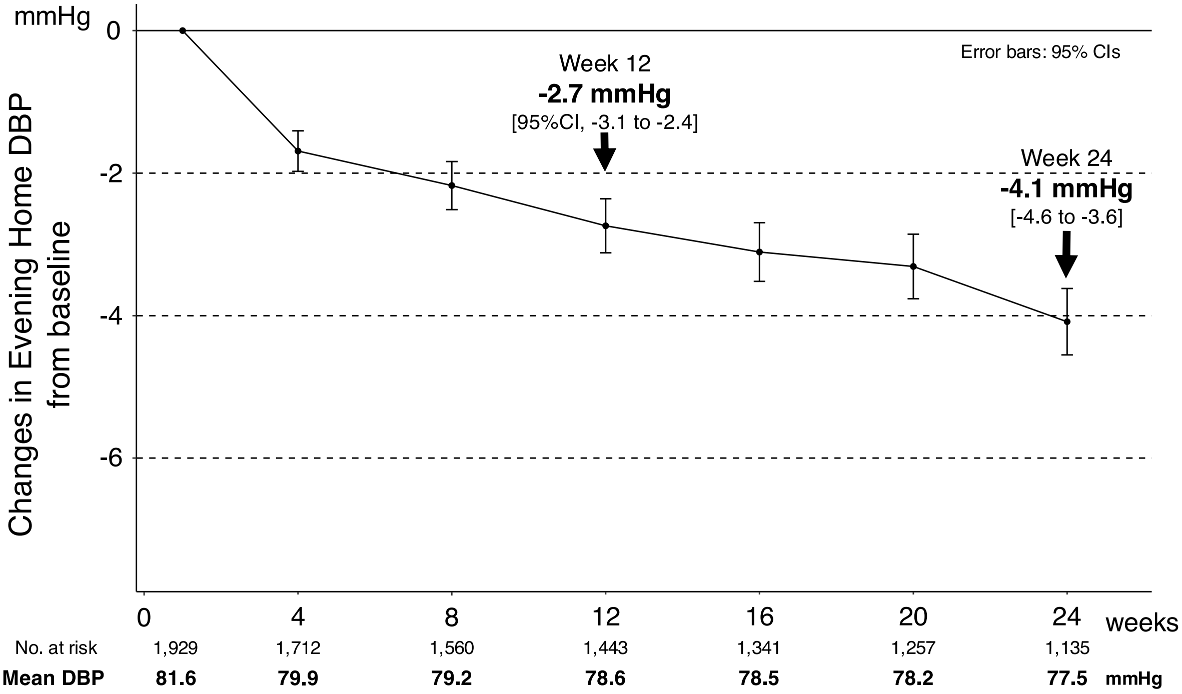


Abbreviations: CI, confidence interval; DBP, diastolic blood pressure.

**Supplementary Figure S4. Achievement rates of blood pressure thresholds according to each hypertension management guideline for morning home blood pressure (A) and evening home blood pressure (B).**

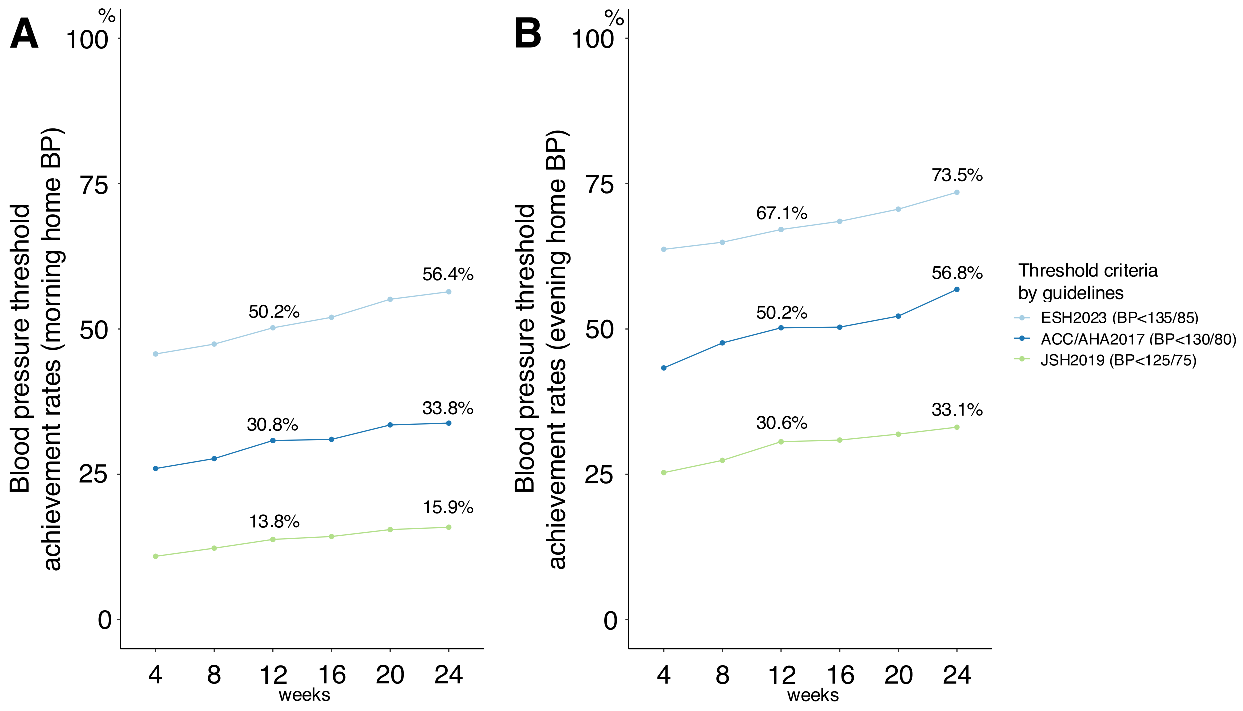


Abbreviations: ACC, American College of Cardiology; AHA, American Heart Association; BP, blood pressure; ESH, European Society of Hypertension; JSH, Japanese Society of Hypertension.

**Supplementary Figure S5. Changes in morning home systolic blood pressure (SBP) from baseline by age (≥65 vs. <65 years).**

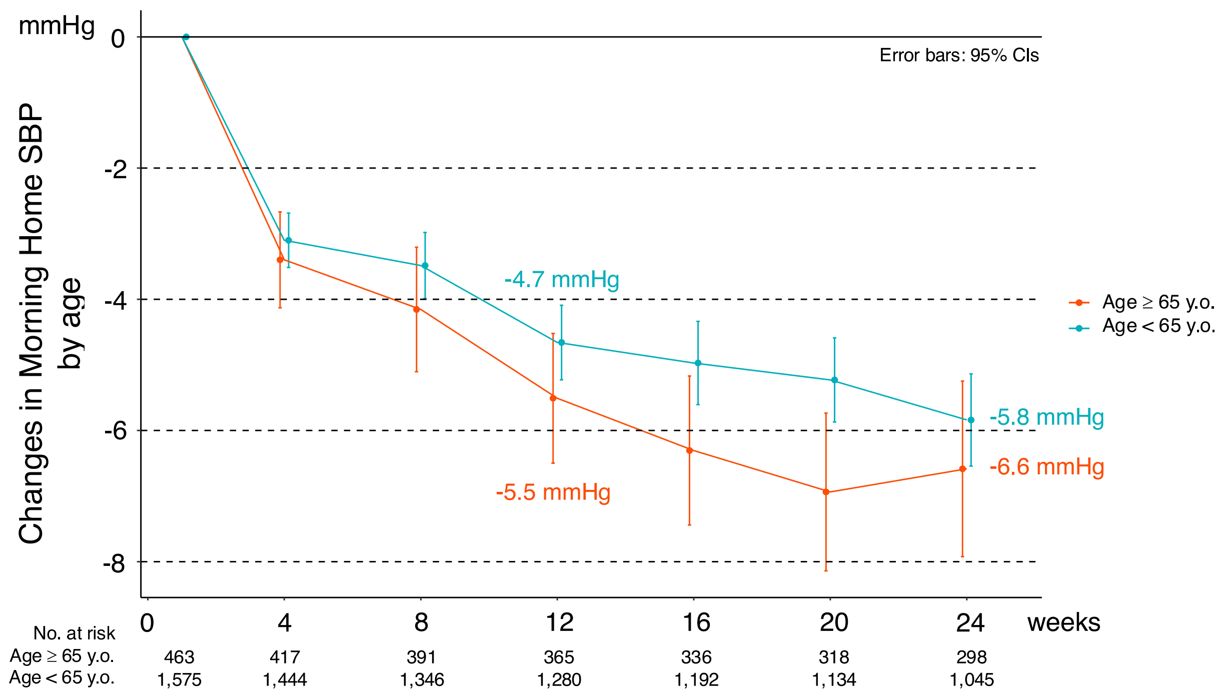

**Supplementary Figure S6. Changes in morning home systolic blood pressure (SBP) from baseline according to sex (male vs. female).**

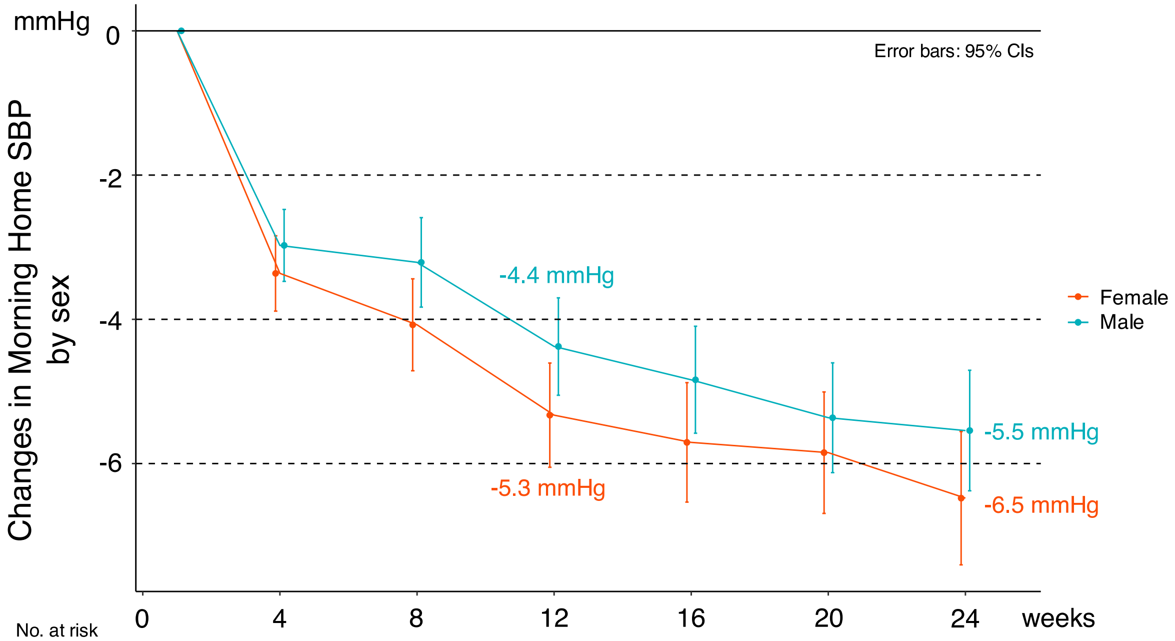


| No. at risk | | | | | | | |
|---|---|---|---|---|---|---|---|
| Male | 1,043 | 939 | 882 | 832 | 778 | 736 | 679 |
| Female | 995 | 922 | 855 | 813 | 750 | 716 | 664 |

**Supplementary Figure S7. Changes in morning home systolic blood pressure (SBP) from baseline according to body mass index (BMI, ≥25 vs. <25 kg/m$^2$).**

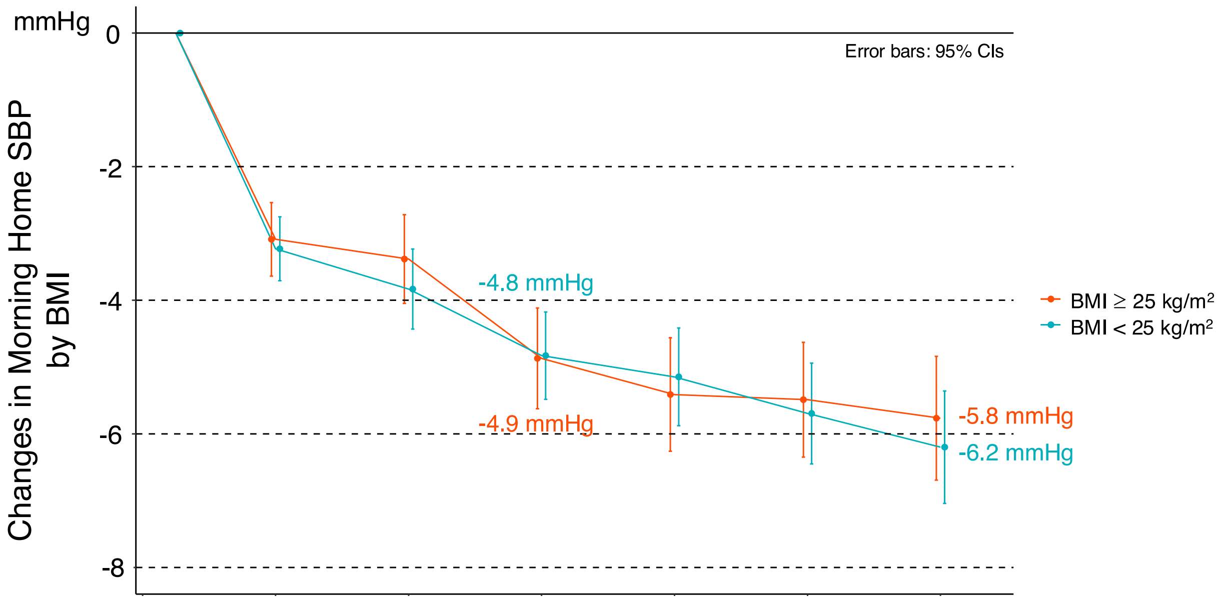


| No. at risk | 0 | 4 | 8 | 12 | 16 | 20 | 24 |
|---|---|---|---|---|---|---|---|
| BMI ≥ 25 | 906 | 821 | 759 | 727 | 678 | 647 | 602 |
| BMI < 25 | 1,132 | 1,040 | 978 | 918 | 850 | 805 | 741 |

**Supplementary Figure S8. Changes in morning home systolic blood pressure (SBP) from baseline according to salt check-sheet score (≥14 vs. <14 points).**

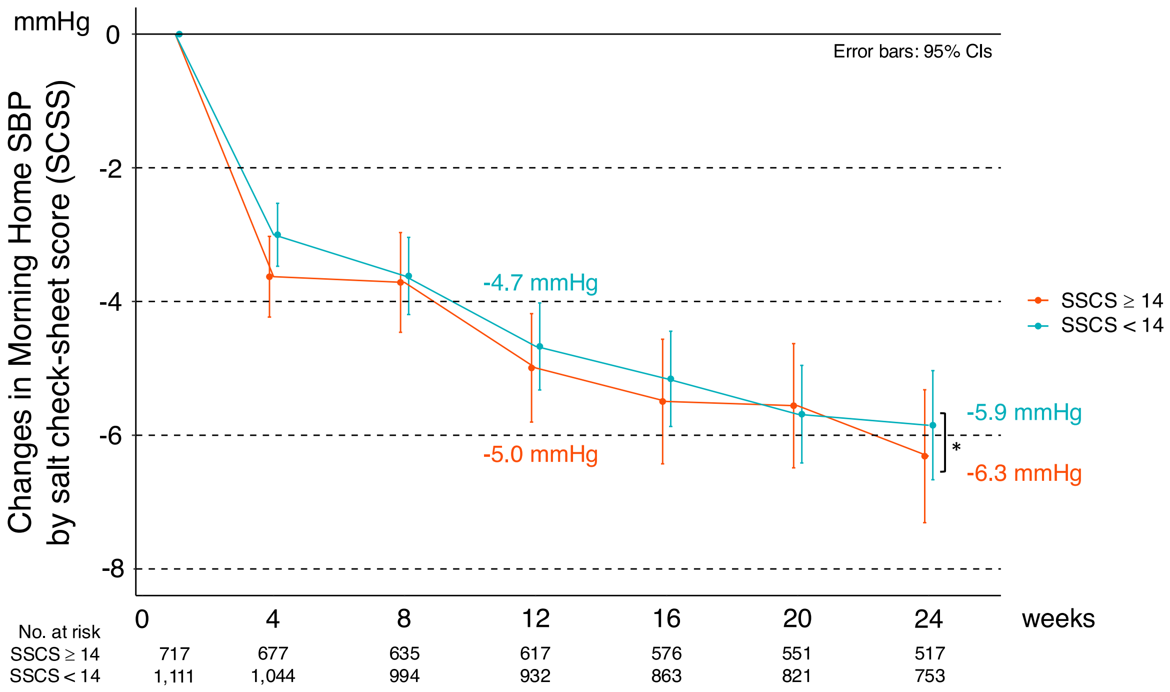

**Supplementary Figure S9. Changes in morning home systolic blood pressure (SBP) from baseline according to medication status (medication[+], no medication, or unknown).**

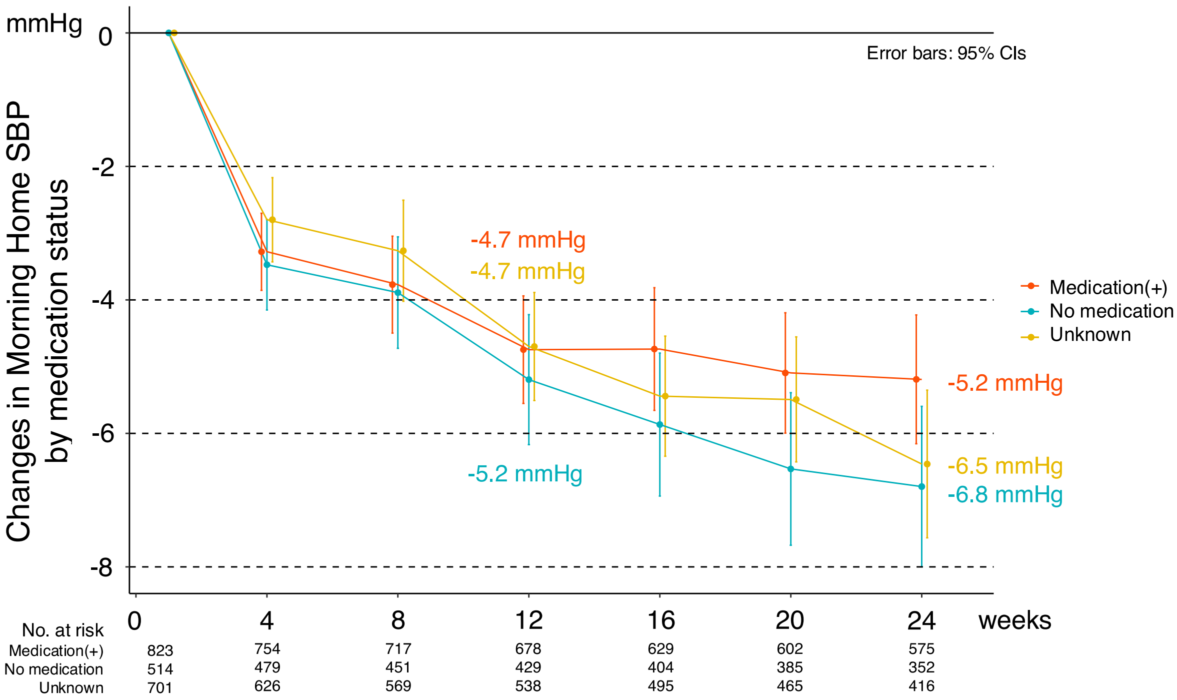

**Supplementary Figure S10. Transitions in morning home systolic blood pressure (SBP) according to baseline SBP levels.**

The extent of SBP reduction was significantly influenced by baseline morning home SBP levels: DTx users with higher baseline morning home SBP exhibited greater reductions in SBP levels from baseline.

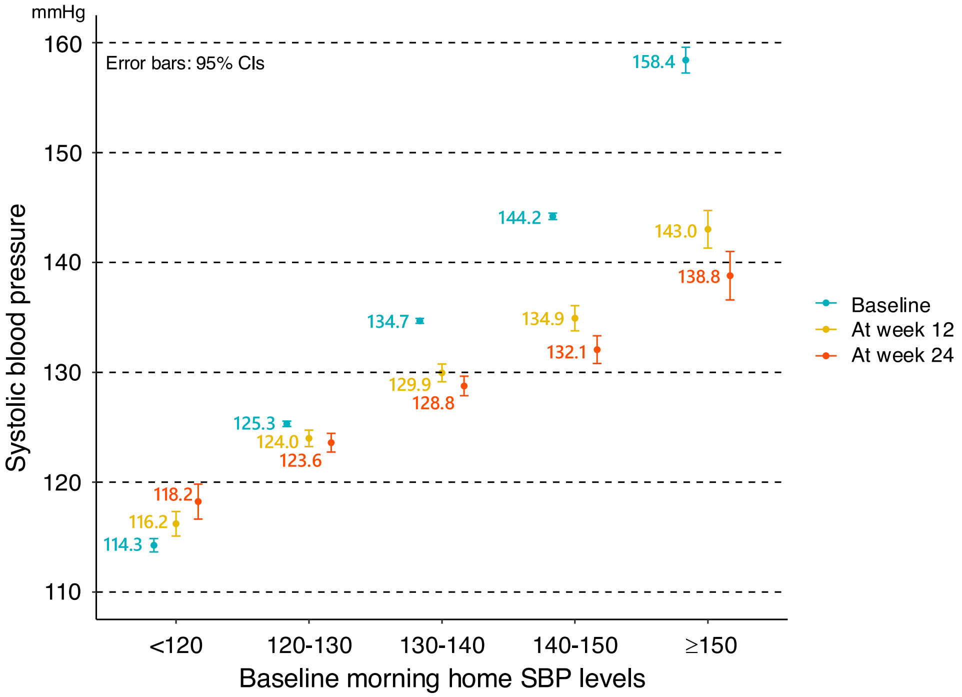


Abbreviations: DTx, digital therapeutics; SBP, systolic blood pressure.

**Supplementary Figure S11. Correlations between morning home SBPs at baseline, week 12 (A), and week 24 (B).**

Intersection points between the regression line and line of equality at both weeks 12 and 24 occurred at around 120 mmHg. These points indicated the maximum achievement targets for morning home SBP through comprehensive lifestyle changes using the DTx app for hypertension.

Abbreviations: SBP, systolic blood pressure.

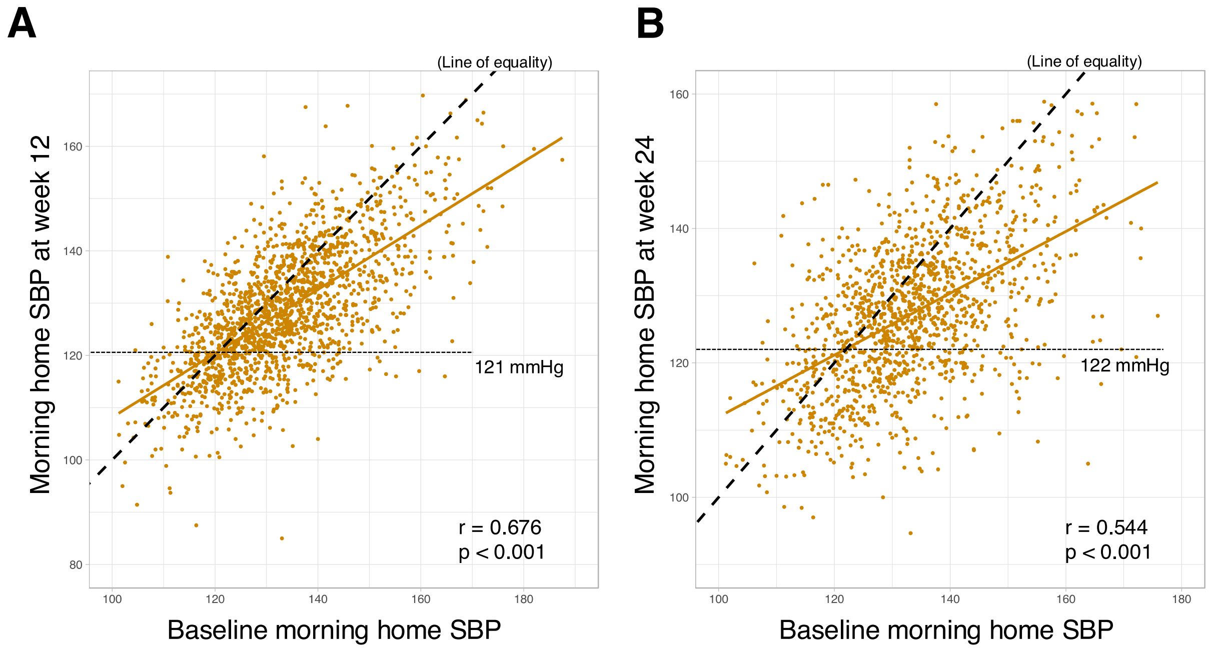

**Supplementary Figure S12. Transitions in morning home SBP at 24 weeks according to baseline morning home SBP levels (A) and season of DTx initiation (B).**

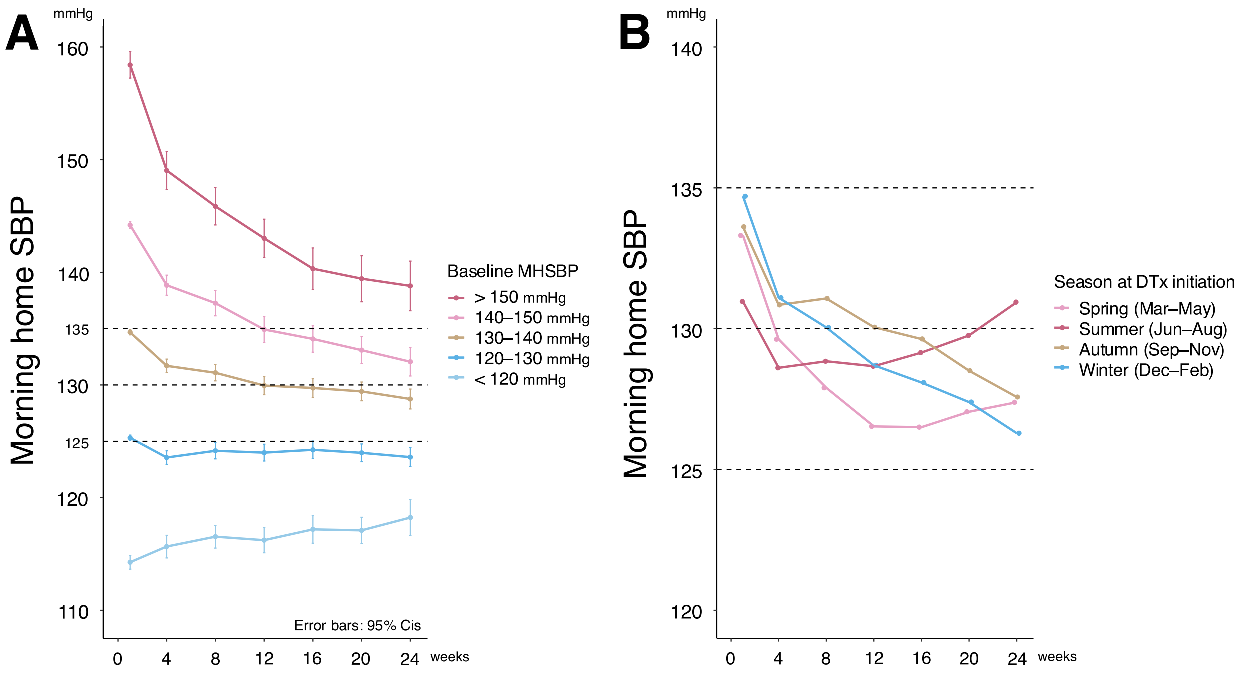


Abbreviations: DTx, digital therapeutics; MHSBP, morning home systolic blood pressure; SBP, systolic blood pressure.
